# Mapping unexplained genetic correlations across 14 psychiatric disorders

**DOI:** 10.64898/2026.01.14.26344089

**Authors:** Yu Chang, Ming-Hong Hsieh, Po-Chung Ju, Cheng-Chen Chang

## Abstract

**Background:** Transdiagnostic genetic factor models organize shared liability across psychiatric disorders, but they may leave systematic pairwise genetic overlap unexplained.

**Methods:** Using publicly available PGC cross-disorder LD score regression genetic correlations and published five-factor genomic SEM parameters, we computed model-implied disorder correlations and derived edge-level residual genetic correlations (observed minus model-implied) for all disorder pairs. We summarized residual misfit by ranking the largest residual edges and by aggregating residual edges into disorder-level hub indices. As a parsimonious comparison, we constructed a p-factor-augmented baseline and repeated the residual mapping. Uncertainty was propagated via Monte Carlo sampling using reported standard errors.

**Results:** Residual structure was concentrated in a subset of disorders rather than being uniformly distributed. The largest positive residual edge was OCD-anxiety (∼0.35), followed by prominent residual links including OCD-Tourette syndrome, ADHD-cannabis use disorder, and ASD-ADHD. At the node level, OCD emerged as the most consistent residual hub, with ADHD typically second. Under the p-factor baseline, the overall residual pattern persisted. Hub rankings did not map one-to-one onto disorder uniqueness, indicating complementary information captured by node-level and edge-level residuals.

**Conclusions:** Higher-order genetic factors explain broad shared liability but leave meaningful, structured residual links between specific disorder pairs. OCD and ADHD are robust residual hubs, highlighting candidate cross-disorder connections for targeted phenotypic harmonization, cross-phenotype GWAS, and theory-guided model refinements.

## Introduction

Transdiagnostic approaches to psychiatric nosology have advanced over recent decades. Frameworks such as the Research Domain Criteria (RDoC) ^1,2^, dimensional extensions of the Diagnostic and Statistical Manual of Mental Disorders (DSM) ^3^, and the Hierarchical Taxonomy of Psychopathology (HiTOP) have encouraged research that moves beyond traditional categorical boundaries ^4,5^. In parallel, genetic research has provided tools for characterizing shared liability across disorders ^6–8^, including general liability models such as the p factor and multifactor structures ^9,10^. Together, these developments have strengthened the evidence for both shared and disorder specific components of psychopathology.

A recent large-scale integration by the Psychiatric Genomics Consortium (PGC) Cross Disorder Working Group synthesized genome wide association study (GWAS) from 14 psychiatric disorders ^11^. Using genomic structural equation modeling, the authors derived five latent genetic factors labeled compulsive, schizophrenia-bipolar (SB), neurodevelopmental, internalizing, and substance use disorder. This model provides a coherent genetic organization of psychiatric disorders and supports the idea that cross disorder risk can be described at a higher dimensional level.

Most transdiagnostic work has focused on establishing classification structures, whereas less attention has been paid to evaluating where such structures succeed and where they leave important patterns unexplained ^12^. In genomic structural equation modeling (SEM) ^13^, model misfit can be examined through residuals ^14^, which capture associations not accounted for by the factor structure. Importantly, residuals can be considered at two levels that address different scientific questions ^15^. Node level residual variance, also referred to as uniqueness, quantifies the proportion of a disorder’s genetic variance that is not explained by the shared factors and therefore highlights disorder specific genetic liability. Edge level residual association focuses on pairs of disorders. It evaluates whether the genetic correlation (rg) between two disorders is larger or smaller than what is implied by the factor model, thereby highlighting cross domain links and boundary tensions that are not captured by the higher order structure.

In the present study, we conduct a secondary analysis using published summary-level results within the five-factor genomic structural equation modeling framework. Our aim is to characterize model misfit at both the edge and node levels by estimating residual pairwise genetic correlations for all disorder pairs and summarizing how residual deviations concentrate across disorders. We rank residual genetic correlations to identify disorder pairs with the largest unexplained links, and aggregate residual edges into disorder-level hub indices to summarize overall residual connectivity across disorders ^16^. Our goal is to contribute a complementary lens on psychiatric nosology by identifying disorder pairs whose genetic correlations remain unexplained by higher-order structures.

## Methods

### 1. Data source from the PGC Cross Disorder study

We used publicly available summary results from the Psychiatric Genomics Consortium Cross Disorder Working Group. The original study integrated GWAS results from 14 psychiatric disorders in a sample of roughly one million individuals. The study first estimated pairwise genetic correlations across disorders using linkage disequilibrium (LD) score regression ^17^. It then used genomic structural equation modeling to derive a latent factor structure for shared genetic liability ^18^. In the original study, exploratory factor analysis was conducted using odd numbered autosomes and confirmatory factor analysis was conducted using even numbered autosomes. This procedure supported a five-factor model. The confirmatory model reported standardized factor loadings, factor correlations, and disorder specific residual variances. It quantifies the proportion of genetic variance that is not explained by the shared factors as a node-level residual measure.

For the present secondary analysis, we extracted the genetic correlation matrix from LD score regression and the standardized confirmatory factor analysis parameter estimates from the supplementary materials.

### 2. Model implied genetic correlations and edge level residuals

For the correlated five-factor model, we computed the model-implied correlation matrix as:

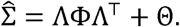

Here, Λ is the matrix of standardized factor loadings. Cross loadings were allowed. Φ is the factor correlation matrix, with ones on the diagonal. Θ is the residual variance matrix for disorders. We placed the standardized residual variance estimates on the diagonal of Θ and set all off diagonal elements to zero.

We then computed the residual genetic correlation matrix as

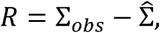

where Σ*_obs_* is the observed genetic correlation matrix from LD score regression. Each element *R^ij^* represents the residual genetic correlation between disorder i and disorder *j* that is not captured by the five-factor structure. Positive values indicate that the observed genetic correlation is larger than the model implied value. Negative values indicate that the observed genetic correlation is smaller than the model implied value.

### 3. P-factor baseline and misfit comparison

In addition to the correlated five-factor model, we evaluated a hierarchical p-factor baseline to assess whether edge-level residual structure persists under a more parsimonious model of general liability. Following the conceptual structure of the original report, we treated covariance among the first-order factors as being mediated by a second-order p factor. Because a complete parameter table for the hierarchical p-factor model was not available in the supplementary spreadsheet, we operationalized this baseline by fitting a one-factor model to the first-order factor correlation matrix Φ, yielding p-factor loadings γ and diagonal residual variances Ψ. We then computed the p-factor-implied disorder correlation matrix:

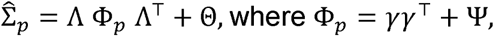

and the corresponding residual matrix 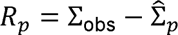. This operational construction is interpreted as a p-factor-augmented baseline for comparing residual structure.

### 4. Statistical analysis

#### 4.1 Edge level residual mapping

We summarized edge level misfit using the residual genetic correlations *R_ij_.* We reported the signed residual values to preserve direction. We ranked disorder pairs by the absolute value | *R_ij_* | to identify the strongest unexplained links.

### 4.2 Residual network hubs

We treated disorders as nodes and residual genetic correlations as weighted edges. We computed several hub indices for each disorder *i* based on the set of residual edges *R_ij_*. We used multiple hub indices because residual edges can differ in magnitude and direction. Strength summarizes the total amount of unexplained connectivity of a disorder across all pairs ^19^. Expected influence preserves edge signs and therefore distinguishes disorders that show predominantly positive versus predominantly negative residual links ^20^. We further decomposed strength into positive and negative components to separate excess cross disorder coupling that is stronger than implied by the factor model from relationships that are weaker than implied by the model.

Strength was defined as

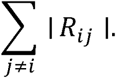

Expected influence was defined as

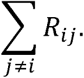

Positive strength was defined as

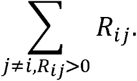

Negative strength was defined as

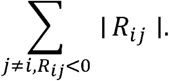

We used these indices to rank disorders and identify the top hub disorders in the residual network.

### 4.3 Uncertainty propagation by Monte Carlo sampling

We did not have access to the full sampling covariance structure for the observed genetic correlations, or the joint sampling covariance across SEM parameters. We therefore propagated uncertainty using Monte Carlo sampling based on reported standard errors (SE). In each simulation, we sampled observed genetic correlations on the Fisher z scale to improve numerical stability. Specifically, each correlation was transformed using *z* = atanh (*r*), and its standard error on the z scale was approximated via the delta method, *SE* ≈ *SE_r_*/(1 - *r*^2^). We then sampled *z* from a normal distribution centered at the transformed estimate and transformed back using *r* = *tanh* (*z*). Sampled correlations were truncated to [-0.99,0.99], assembled into a symmetric observed correlation matrix with unit diagonal, and then projected to the nearest valid correlation matrix to ensure a positive semidefinite structure ^21^.

We also sampled standardized confirmatory factor model parameters from approximately normal distributions using their reported standard errors. These parameters included factor loadings, factor correlations, and disorder-specific residual variances. For each draw, we reconstructed the model-implied disorder covariance matrix as 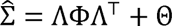 (with Θ diagonal), symmetrized it, and converted it to a model-implied correlation matrix by standardization, 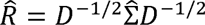, where 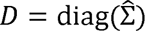. We set the diagonal of the residual matrix to zero and computed residual genetic correlations as *R* = *Σ_Obs_* - *R̂*, where Σ_Obs_ denotes the projected observed correlation matrix in that draw. Because the dependence structure among observed correlations and SEM parameters was unavailable, we treated draws as independent across disorder pairs and SEM parameters; this independence assumption can underestimate uncertainty. We used 3000 simulations with a fixed random seed.

### 4.4 Uncertainty summaries, rank stability, and sensitivity analyses

We summarized uncertainty in edge and node quantities using the Monte Carlo sampling distributions. For node hub indices, we reported the mean and percentile based 95 percent intervals across draws. We evaluated hub ranking stability using a top K (K = 3) inclusion probability ^22^. For each draw, we ranked disorders by node strength in descending order and computed, for each disorder, the proportion of draws in which its rank was within the top three. Ties were handled using average ranks.

To assess sensitivity to potential underestimation of uncertainty under the independence approximation, we repeated the full procedure after inflating standard errors by fixed multipliers, including 1.5 and 2.0, and reevaluated hub summaries and top three inclusion probabilities under these more conservative settings.

### 4.5 Comparison with node level uniqueness

We compared disorder hub indices from the residual network with the disorder uniqueness estimates reported in the confirmatory factor model. We compared node level uniqueness and edge level hub rankings descriptively. For each disorder, we reported its uniqueness rank and residual network strength rank.

## Results

### Five-factor residual network

Visual inspection of the residual correlation matrices indicated that deviations were not uniformly distributed across disorders, with prominent residual structure evident in the residual heatmap (Figure 1) and a distribution centered near zero but with nontrivial tails (Figure 2). The largest positive residuals were concentrated around obsessive-compulsive disorder (OCD) and attention-deficit/hyperactivity disorder (ADHD). The strongest residual edge was observed between OCD and anxiety (ANX), with a residual genetic correlation of 0.35 (95% Confidence interval (CI) 0.26, 0.43). Additional prominent positive residuals included OCD with Tourette syndrome (TS) at 0.29 (0.16, 0.41) and OCD with autism spectrum disorder (ASD) at 0.20 (0.08, 0.32). ADHD showed elevated residual connectivity with cannabis use disorder (CUD), 0.26 (0.17, 0.35), and with post-traumatic stress disorder (PTSD), 0.16 (0.08, 0.23), while ASD and ADHD also exhibited a sizable positive residual association of 0.23 (0.11, 0.35). In contrast, several residual associations were negative, indicating observed genetic correlations lower than those implied by the five-factor structure; these included anorexia nervosa (AN) with ADHD at −0.15 (−0.22, −0.07) and schizophrenia (SCZ) with ADHD at −0.08 (−0.13, −0.02). The top 10 residual genetic correlations by magnitude are summarized in Table 1. (Figures 1-2)

**Figure 1.**
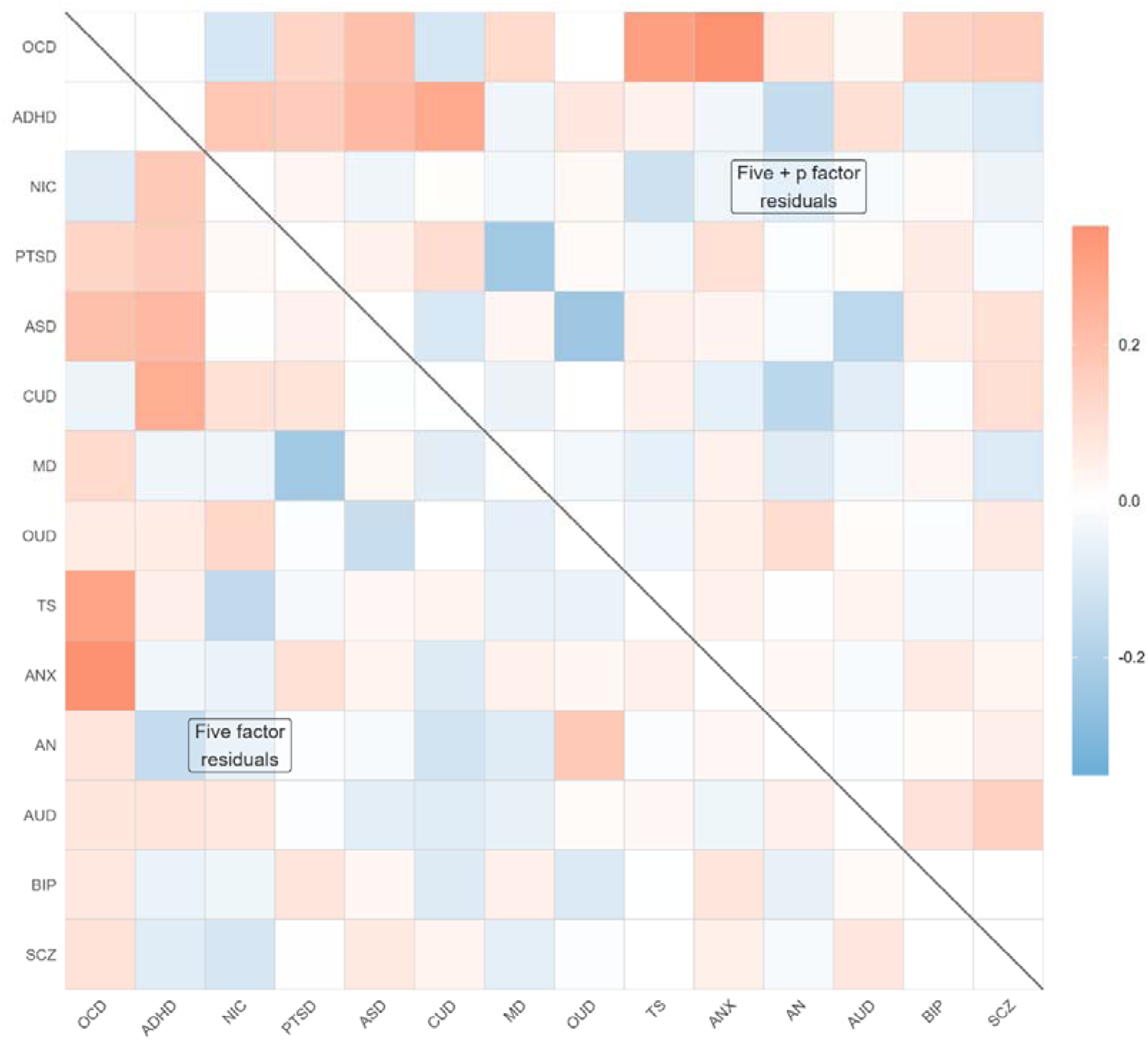
Heatmap of residual genetic correlations (five-factor vs five-factor + p). OCD: obsessive-compulsive disorder; ADHD: attention-deficit/hyperactivity disorder; NIC: nicotine dependence; PTSD: post-traumatic stress disorder; ASD: autism spectrum disorder; CUD: cannabis use disorder; MD: major depression; OUD: opioid use disorder; TS: Tourette syndrome; ANX: anxiety; AN: anorexia nervosa; AUD: alcohol use disorder; BIP: bipolar disorder; SCZ: schizophrenia

**Figure 2.**
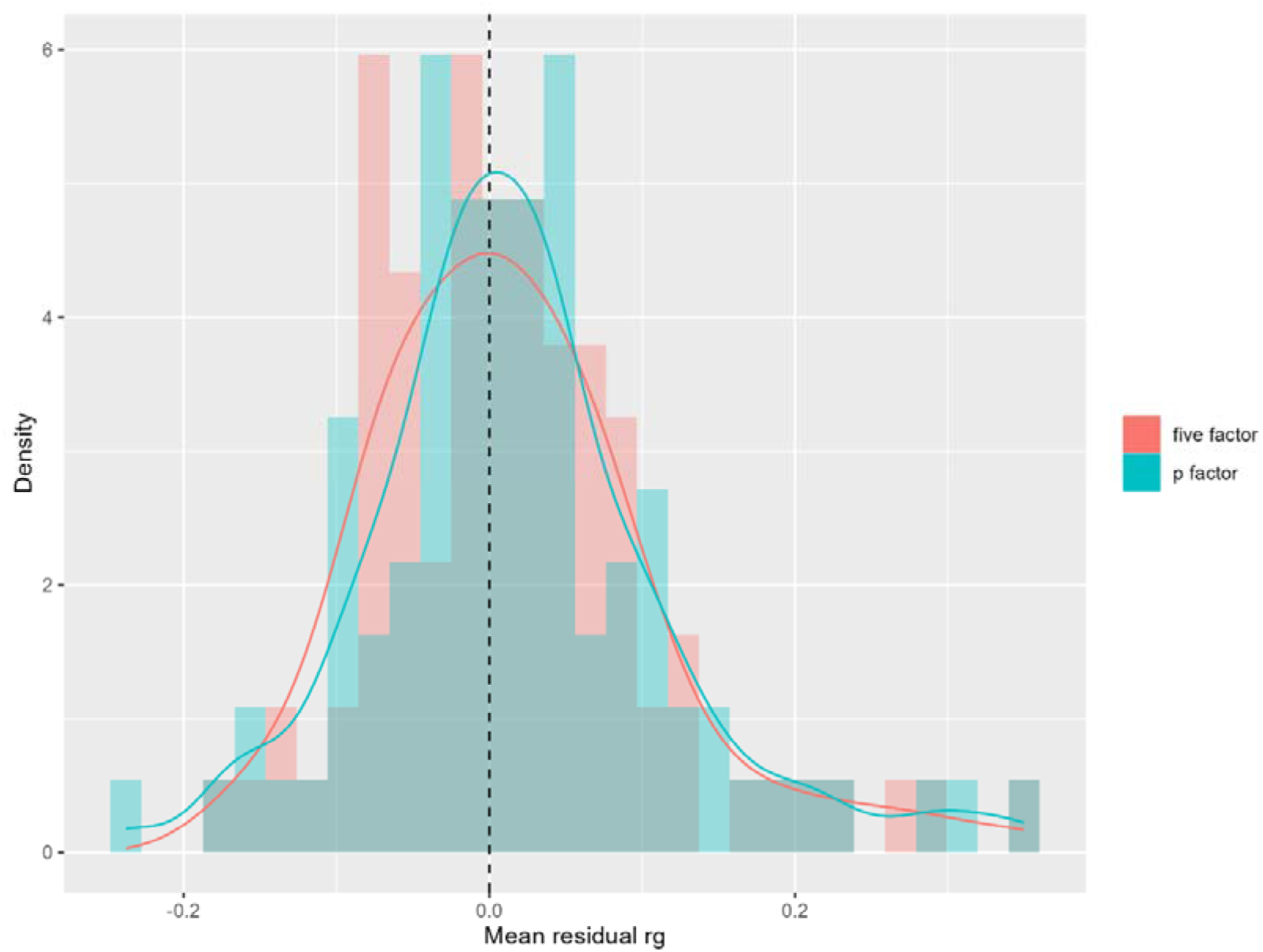
Distribution of mean residual genetic correlations under the five-factor and p-factor models. Note: Dashed line indicates zero. Solid lines show kernel density estimates (KDE). OCD: obsessive-compulsive disorder; ADHD: attention-deficit/hyperactivity disorder; NIC: nicotine dependence; PTSD: post-traumatic stress disorder; ASD: autism spectrum disorder; CUD: cannabis use disorder; MD: major depression; OUD: opioid use disorder; TS: Tourette syndrome; ANX: anxiety; AN: anorexia nervosa; AUD: alcohol use disorder; BIP: bipolar disorder; SCZ: schizophrenia

**Table 1.**
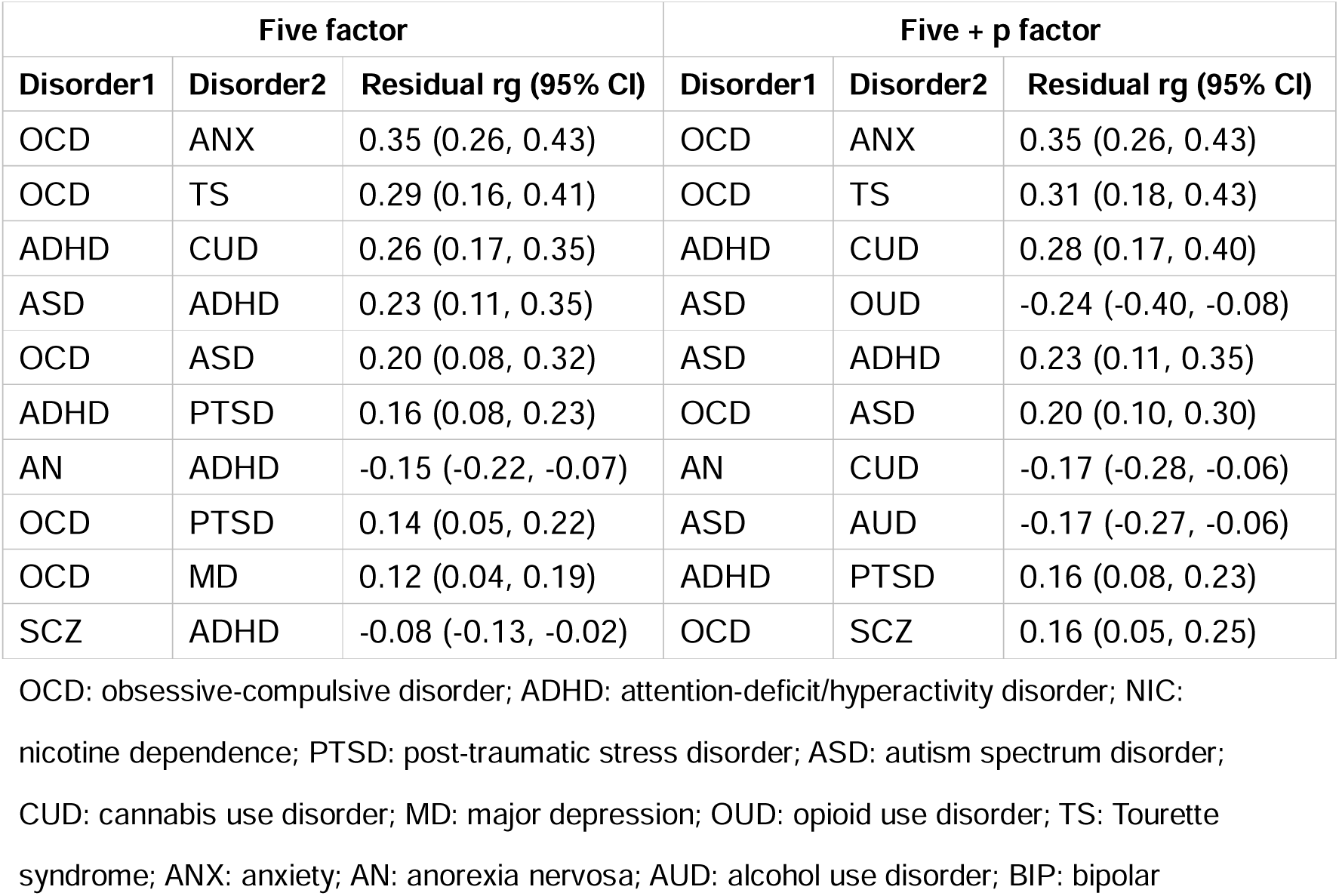
Top 10 residual genetic correlations under the five-factor and five-factor + p models.

| Five factor |  |  | Five + p factor |  |  |
| --- | --- | --- | --- | --- | --- |
| Disorder1 | Disorder2 | Residual rg (95% CI) | Disorder1 | Disorder2 | Residual rg (95% CI) |
| OCD | ANX | 0.35 (0.26, 0.43) | OCD | ANX | 0.35 (0.26, 0.43) |
| OCD | TS | 0.29 (0.16, 0.41) | OCD | TS | 0.31 (0.18, 0.43) |
| ADHD | CUD | 0.26 (0.17, 0.35) | ADHD | CUD | 0.28 (0.17, 0.40) |
| ASD | ADHD | 0.23 (0.11, 0.35) | ASD | ODU | -0.24 (-0.40, -0.08) |
| OCD | ASD | 0.20 (0.08, 0.32) | ASD | ADHD | 0.23 (0.11, 0.35) |
| ADHD | PTSD | 0.16 (0.08, 0.23) | OCD | ASD | 0.20 (0.10, 0.30) |
| AN | ADHD | -0.15 (-0.22, -0.07) | AN | CUD | -0.17 (-0.28, -0.06) |
| OCD | PTSD | 0.14 (0.05, 0.22) | ASD | AUD | -0.17 (-0.27, -0.06) |
| OCD | MD | 0.12 (0.04, 0.19) | ADHD | PTSD | 0.16 (0.08, 0.23) |
| SCZ | ADHD | -0.08 (-0.13, -0.02) | OCD | SCZ | 0.16 (0.05, 0.25) |

OCD: obsessive-compulsive disorder; ADHD: attention-deficit/hyperactivity disorder; NIC: nicotine dependence; PTSD: post-traumatic stress disorder; ASD: autism spectrum disorder; CUD: cannabis use disorder; MD: major depression; OUD: opioid use disorder; TS: Tourette syndrome; ANX: anxiety; AN: anorexia nervosa; AUD: alcohol use disorder; BIP: bipolar

At the node level, hub structure in the five-factor residual network was summarized using strength, defined as the sum of absolute residual edges incident on each disorder. OCD was the most central node (strength 1.71; 95% CI 1.35, 2.08), followed by ADHD (1.44; 1.13, 1.76) and nicotine dependence (NIC) (1.30; 0.84, 1.82), with PTSD (1.26; 0.74, 2.78) and ASD (1.21; 0.81, 1.72) forming the next tier of hubs. Rank stability analyses supported the prominence of the leading hubs, with top-3 inclusion probabilities of 0.96 for OCD and 0.62 for ADHD, and lower probabilities for NIC (0.34) and PTSD (0.31) (Table 3). Sensitivity analyses inflating standard errors by 1.5× and 2.0× widened uncertainty intervals but did not change the qualitative hub pattern; the top-3 hub ordering by strength remained consistent.

**Table 3.** Node-level hub indices in the five-factor residual network.

| Five factor |  |  | Five factor + p |  |  | Uniqueness |  |
| --- | --- | --- | --- | --- | --- | --- | --- |
| Node | Strength (95% CI) | P of top 3 | Node | Strength five plus p (95% CI) | P of top 3 | Node | Estimate (SE) |
| OCD | 1.71(1.35,2.08) | 0.96 | OCD | 1.86(1.52,2.21) | 0.97 | TS | 0.89(0.11) |
| ADHD | 1.44(1.13,1.76) | 0.62 | ADHD | 1.51(1.20,1.84) | 0.54 | ASD | 0.78(0.12) |
| NIC | 1.30(0.84,1.82) | 0.34 | ASD | 1.48(1.11,1.87) | 0.47 | NIC | 0.62(0.15) |
| PTSD | 1.26(0.74,2.78) | 0.31 | PTSD | 1.48(0.85,2.47) | 0.44 | ADHD | 0.59(0.04) |
| ASD | 1.21(0.81,1.72) | 0.18 | CUD | 1.34(0.98,1.72) | 0.19 | OCD | 0.52(0.09) |
| CUD | 1.21(0.88,1.57) | 0.16 | MD | 1.33(0.70,2.32) | 0.27 | AN | 0.52(0.08) |
| MD | 1.17(0.66,2.73) | 0.21 | OD | 1.14(0.76,1.57) | 0.05 | SCZ | 0.32(0.04) |
| OD | 1.14(0.76,1.56) | 0.12 | NIC | 1.13(0.74,1.56) | 0.04 | BIP | 0.32(0.04) |
| TS | 1.09(0.75,1.45) | 0.05 | TS | 1.10(0.77,1.47) | 0.02 | CUD | 0.31(0.10) |
| ANX | 1.07(0.81,1.34) | 0.02 | ANX | 1.05(0.78,1.34) | 0.00 | ANX | 0.28(0.05) |
| AN | 1.03(0.72,1.35) | 0.02 | SCZ | 1.04(0.74,1.35) | 0.00 | AUD | 0.28(0.06) |
| AUD | 0.83(0.56,1.11) | 0.00 | AN | 1.02(0.73,1.36) | 0.00 | PTSD | 0.18(0.05) |
| BIP | 0.79(0.55,1.05) | 0.00 | AUD | 0.98(0.68,1.34) | 0.00 | OD | 0.15(0.15) |
| SCZ | 0.76(0.55,0.99) | 0.00 | BIP | 0.80(0.53,1.10) | 0.00 | MD | 0.01(0.04) |

OCD: obsessive-compulsive disorder; ADHD: attention-deficit/hyperactivity disorder; NIC: nicotine dependence; PTSD: post-traumatic stress disorder; ASD: autism spectrum disorder; CUD: cannabis use disorder; MD: major depression; OD: opioid use disorder; TS: Tourette syndrome; ANX: anxiety; AN: anorexia nervosa; AUD: alcohol use disorder; BIP: bipolar disorder; SCZ: schizophrenia

Signed indices clarified the directionality underlying these hubs. Expected influence (EI) was strongly positive for OCD (EI 1.37; 95% CI 0.85, 1.82) and ADHD (0.67; 0.29, 1.03), consistent with strength being predominantly driven by positive residual edges (OCD Spos 1.54 vs Sneg 0.17; ADHD Spos 1.05 vs Sneg 0.38). In contrast, NIC and major depression (MD) showed EI near zero or negative (NIC −0.05; −0.93, 0.81; MD −0.48; −2.21, 0.20), reflecting substantial counterbalancing of positive and negative residual connectivity (NIC Spos 0.63 vs Sneg 0.67; MD Spos 0.35 vs Sneg 0.82). Full node-level hub indices (strength, EI, Spos, and Sneg) for the five-factor residual network are shown in Table 4.

**Table 4.** Node-level hub indices in the five-factor residual network.

| Node | Strength | EI | Spos | Sneg |
| --- | --- | --- | --- | --- |
| **OCD** | 1.71(1.35,2.08) | 1.37(0.85,1.82) | 1.54(1.15,1.92) | 0.17(0.01,0.39) |
| **ADHD** | 1.44(1.13,1.76) | 0.67(0.29,1.03) | 1.05(0.75,1.35) | 0.38(0.22,0.55) |
| **NIC** | 1.30(0.84,1.82) | -0.05(-0.93,0.81) | 0.63(0.20,1.17) | 0.67(0.25,1.23) |
| **PTSD** | 1.26(0.74,2.78) | 0.34(-1.39,1.04) | 0.80(0.52,1.12) | 0.46(0.02,2.04) |
| **ASD** | 1.21(0.81,1.72) | 0.42(-0.36,1.17) | 0.82(0.40,1.36) | 0.40(0.07,0.85) |
| **CUD** | 1.21(0.88,1.57) | 0.03(-0.50,0.55) | 0.62(0.34,0.95) | 0.59(0.29,0.94) |
| **MD** | 1.17(0.66,2.73) | -0.48(-2.21,0.20) | 0.35(0.17,0.57) | 0.82(0.29,2.45) |
| **OUD** | 1.14(0.76,1.56) | 0.11(-0.51,0.73) | 0.63(0.27,1.04) | 0.51(0.19,0.88) |
| **TS** | 1.09(0.75,1.45) | 0.18(-0.53,0.90) | 0.63(0.30,1.11) | 0.45(0.11,0.89) |
| **ANX** | 1.07(0.81,1.34) | 0.54(0.18,0.92) | 0.81(0.58,1.08) | 0.27(0.08,0.49) |
| **AN** | 1.03(0.72,1.35) | -0.20(-0.68,0.27) | 0.41(0.18,0.69) | 0.61(0.32,0.96) |
| **AUD** | 0.83(0.56,1.11) | 0.16(-0.29,0.60) | 0.49(0.25,0.79) | 0.33(0.11,0.60) |
| **BIP** | 0.79(0.55,1.05) | 0.01(-0.38,0.39) | 0.40(0.20,0.63) | 0.39(0.17,0.64) |
| **SCZ** | 0.76(0.55,0.99) | 0.04(-0.33,0.42) | 0.40(0.20,0.65) | 0.36(0.16,0.59) |

EI: Expected Influence; Spos: positive strength; Sneg: negative strength. OCD: obsessive-compulsive disorder; ADHD: attention-deficit/hyperactivity disorder; NIC: nicotine dependence; PTSD: post-traumatic stress disorder; ASD: autism spectrum disorder; CUD: cannabis use disorder; MD: major depression; OUD: opioid use disorder; TS: Tourette syndrome; ANX: anxiety; AN: anorexia nervosa; AUD: alcohol use disorder; BIP: bipolar disorder; SCZ: schizophrenia

### Five-factor + p-factor residual network

We next repeated the residual analysis under the p-factor-augmented baseline. The overall pattern of residuals remained centered near zero with a similar distributional shape, as shown by the density plots of mean residual genetic correlations (Figure 2), and the residual heatmap continued to show concentrated residual structure in a subset of disorders (Figure 1). At the edge level, the largest residual remained OCD-ANX at 0.35 (0.26, 0.43), and OCD-TS remained elevated at 0.31 (0.18, 0.43). ADHD continued to show a strong positive residual association with CUD at 0.28 (0.17, 0.40), and ASD-ADHD remained sizable at 0.23 (0.11, 0.35). The p-factor baseline additionally highlighted negative residual associations between ASD and opioid use disorder (OUD), −0.24 (−0.40, −0.08), and between ASD and alcohol use disorder (AUD), −0.17 (−0.27, −0.06), alongside a negative residual between AN and CUD, −0.17 (−0.28, −0.06). A positive residual edge between OCD and SCZ was also observed at 0.16 (0.05, 0.25). The top 10 residual genetic correlations by magnitude under this baseline are summarized in Table 1. (Figures 1-2)

Node-level hub rankings under the p-factor-augmented baseline retained OCD as the dominant residual hub (strength 1.86; 95% CI 1.52, 2.21) and placed ADHD second (1.51; 1.20, 1.84), with ASD increasing in centrality to become the third-ranked hub (1.48; 1.11, 1.87). Rank stability estimates mirrored this pattern, with top-3 inclusion probabilities of 0.97 for OCD, 0.54 for ADHD, and 0.47 for ASD. These node-level strength estimates and rank stability statistics are summarized in Table 3. Results were robust to 1.5× and 2.0×standard-error inflation, with the top-3 hub ordering by strength remaining consistent.

Signed indices indicated that OCD and ADHD continued to show clearly positive net residual connectivity (EI 1.33; 95% CI 0.86, 1.80 and 0.70; 0.28, 1.11, respectively), whereas ASD’s net influence was smaller despite high strength (EI 0.17; −0.38, 0.73), consistent with a mixture of positive and negative residual edges (ASD Spos 0.83 vs Sneg 0.66). NIC remained characterized by comparatively larger negative components (EI −0.25; −0.93, 0.44; Spos 0.44 vs Sneg 0.69), and CUD likewise showed EI near zero (−0.03; −0.58, 0.52). Full node-level hub indices (strength, EI, Spos, and Sneg) for the p-factor residual network are shown in Table 5.

**Table 5.**
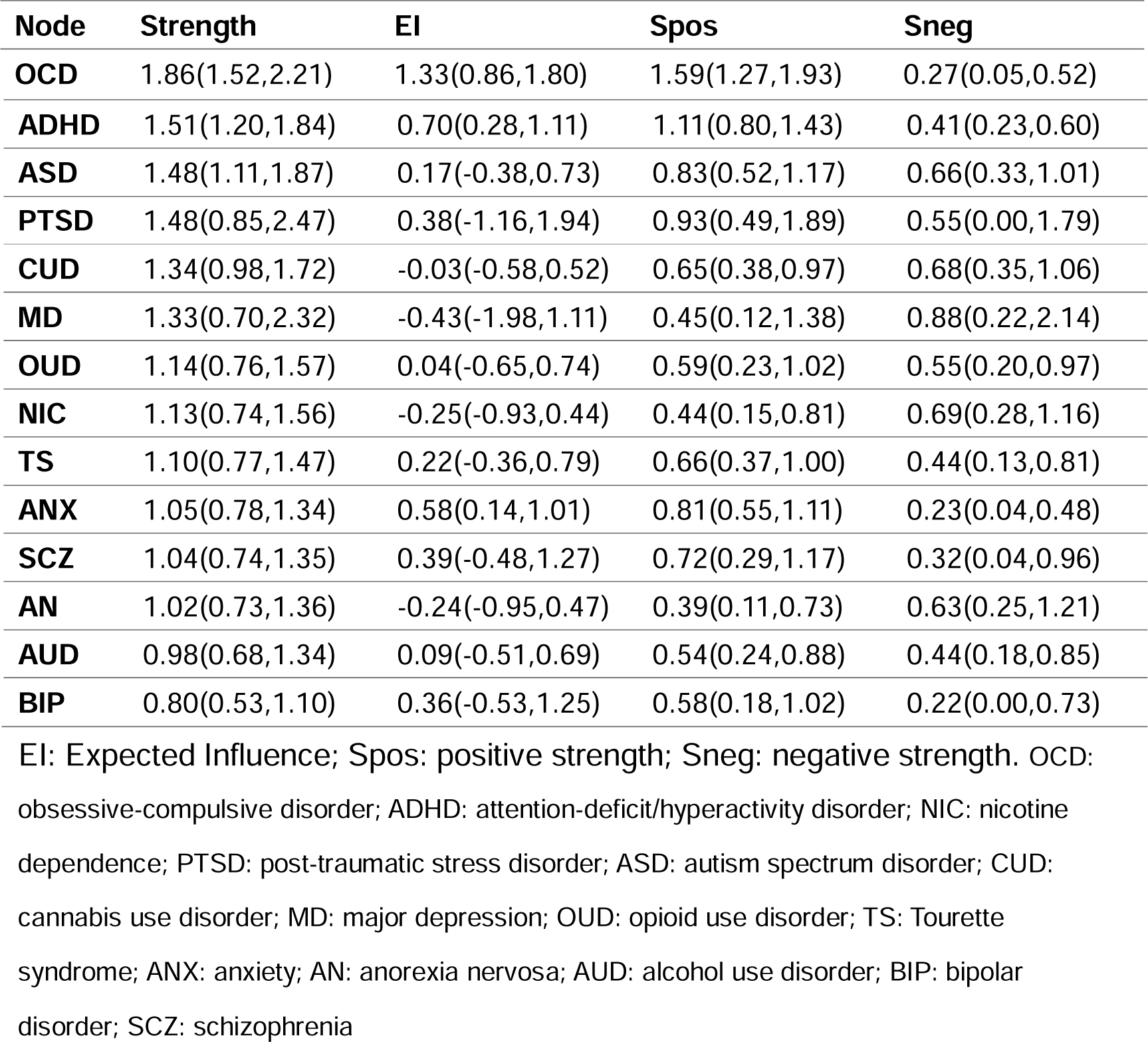
Node-level hub indices in the five-factor + p residual network.

### Uniqueness estimates and association with residual hub structure

Uniqueness varied substantially across disorders, with the highest values observed for TS and ASD and the lowest for MD (Table 3). However, uniqueness did not map one-to-one onto residual hub strength. OCD emerged as the strongest residual hub despite only intermediate uniqueness, indicating that its residual structure is expressed primarily through concentrated pairwise deviations rather than large disorder-specific residual variance. Conversely, TS showed very high uniqueness but comparatively modest residual hub strength, suggesting that its unexplained genetic liability is largely disorder-specific rather than distributed across multiple residual cross-disorder links. ASD combined high uniqueness with high residual hub strength under the p-factor-augmented baseline, reflecting both substantial trait-specific variance and meaningful residual connections that persist beyond general and specific transdiagnostic factors. The strength-uniqueness comparison highlights that node-level and edge-level residuals capture complementary aspects of model departure, a pattern summarized visually in the joint plot of residual hub strength and uniqueness with uncertainty intervals (Figure 3).

**Figure 3.**
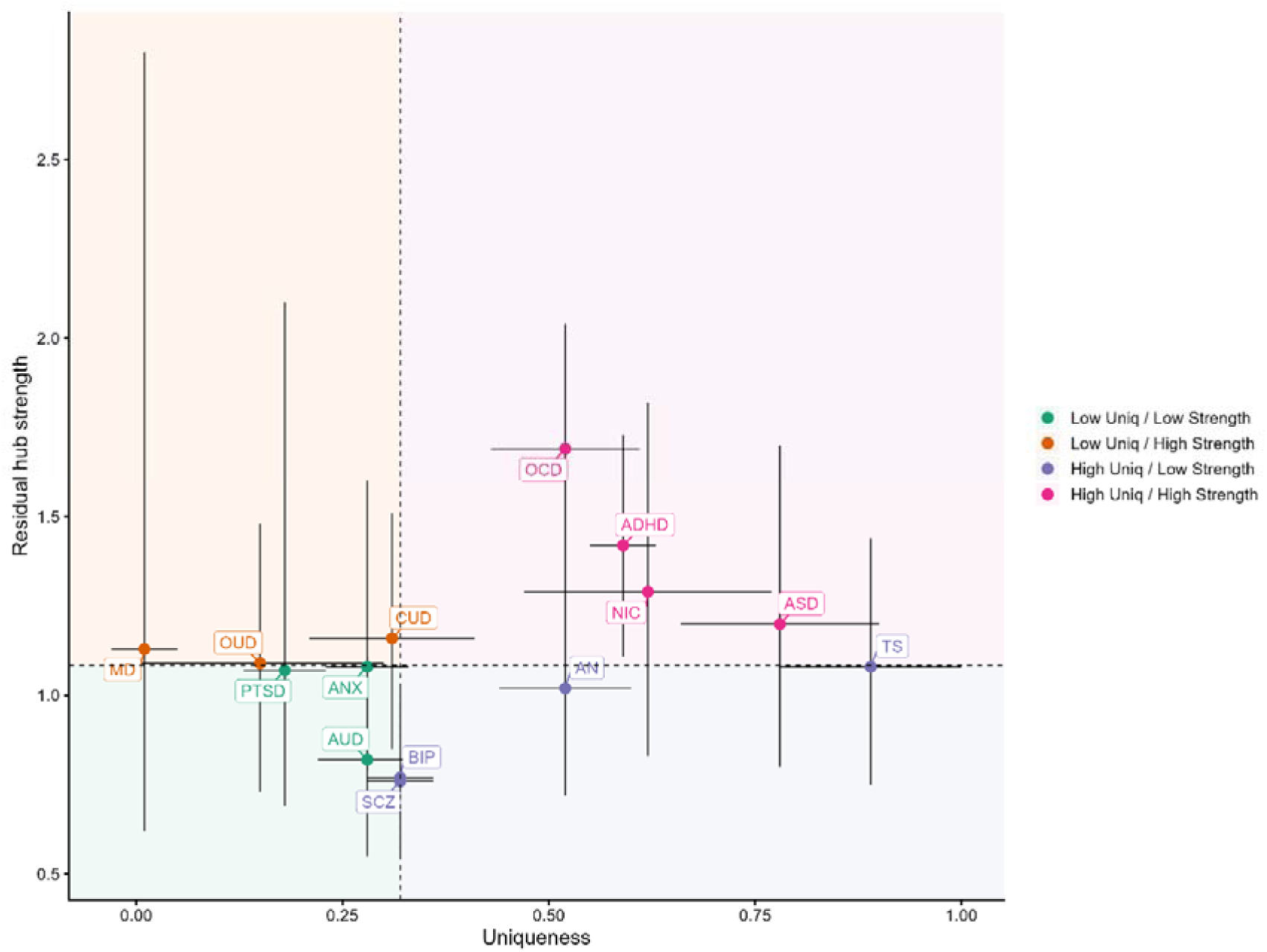
Residual hub strength versus uniqueness in the five-factor residual network. Vertical error bars indicate 95% CIs for strength and horizontal error bars indicate SEs for uniqueness; dashed lines denote median splits. OCD: obsessive-compulsive disorder; ADHD: attention-deficit/hyperactivity disorder; NIC: nicotine dependence; PTSD: post-traumatic stress disorder; ASD: autism spectrum disorder; CUD: cannabis use disorder; MD: major depression; OUD: opioid use disorder; TS: Tourette syndrome; ANX: anxiety; AN: anorexia nervosa; AUD: alcohol use disorder; BIP: bipolar disorder; SCZ: schizophrenia

## Discussion

This study evaluated where a five-factor genetic structure explained cross disorder genetic overlap and where it left unexplained. Residual genetic correlations captured pairwise overlap that remained after accounting for the shared factor structure. The largest residual edges involved OCD, including OCD-ANX as a cross-domain link and OCD-TS within the compulsive domain. Additional prominent residual links included ADHD-CUD and ASD-ADHD within the neurodevelopmental domain. Together, these residual edges highlight both boundary-spanning connections and within-domain ties that were not fully captured by the five-factor model ^23–26^. Clinically, these results nominate specific cross-domain pairings that may warrant proactive assessment and integrated screening even when disorders are placed in distinct higher-order domains.

Node strength captured overall residual connectivity and identified OCD as the dominant hub and ADHD as a consistent second hub across both models. Directional indices refined this picture: OCD and ADHD showed positive expected influence largely driven by positive residual edges, whereas nicotine dependence and major depression showed expected influence near or below zero because positive and negative residual components were more balanced or skewed negative. Predominantly positive hubs may motivate targeted model extensions such as cross-loadings or selected residual covariances, whereas mixed or negative deviations may indicate overgeneralized shared structure and motivate re-specification of loadings, factor correlations, or contrast dimensions.

Adding a general factor preserved the main hub pattern. OCD remained the top hub and ADHD remained second. ASD increased in strength and became the third hub, whereas nicotine dependence decreased in strength and moved to a lower rank. Overall, the p-factor-augmented baseline pulled many residual correlations closer to zero, consistent with general liability absorbing broad shared overlap. At the same time, several residual edges became more pronounced, including negative residual links between ASD and OUD and between ASD and AUD. This pattern suggests that the observed ASD-substance-use overlap is lower than expected under a p-mediated structure and may reflect residual contrasts not captured by general liability.

Uniqueness quantified disorder specific genetic variance that the shared factors did not explain. TS and ASD showed the largest uniqueness estimates. Major depression showed very small uniqueness. These patterns did not match residual hub strength ranks. OCD and ADHD were strong residual hubs but they were not the highest in uniqueness. TS had the highest uniqueness but it was not a top strength hub. This contrast supported the view that uniqueness and residual network structure capture complementary information.

As an external point of reference ^27^, prior work using a clinical comorbidity network estimated via partial correlations reported that the most prominent hubs were concentrated in substance use disorder categories and developmental phenotypes, including intellectual disability nodes. Because partial-correlation networks reflect conditional comorbidity patterns in clinical data, direct correspondence with residual genetic correlations is not expected. In that clinical hub profile, OCD ranked comparatively low despite being the dominant hub in our genetic residual network. At the same time, the prominence of substance-related hubs in the clinical network is broadly consistent with well-known co-occurrence patterns in clinical settings. The centrality of intellectual disability and related developmental phenotypes further suggests that developmental conditions, many of which lack direct counterparts in current cross-disorder GWAS resources, may be important targets for future integrative genetic and clinical network studies.

By quantifying edge-level residual genetic correlations and aggregating them into node-level residual hubs, we provide a principled way to rank where transdiagnostic factor models leave systematic unexplained structure. Residual edges pinpoint specific disorder pairs for which the observed genetic overlap deviates from model expectations, thereby generating concrete, testable targets for refinement. First, they motivate closer scrutiny of phenotype definitions and case ascertainment for the implicated pairs ^28^, including harmonizing diagnostic criteria across cohorts and evaluating whether residual links are driven by comorbidity, symptom heterogeneity, or measurement differences. Second, they nominate priorities for dimensional and cross-phenotype GWAS, such as constructing symptom-based or trait-based measures that bridge the paired disorders and testing whether these dimensional phenotypes capture the residual overlap more directly than binary case definitions. Third, they provide focused candidates for model re-specification, including theory-guided cross-loadings, selective residual covariances, or additional domain-specific factors designed to account for reproducible residual structure ^29^.

Because residual structure may reflect cohort composition, GWAS wave, phenotype definition, and ancestry-related architecture ^30^, the resulting rankings should be treated as hypothesis-generating and prioritized for replication across independent GWAS releases and ancestries before drawing etiological conclusions. Future work can formalize these diagnostics by testing SEM variants that incorporate only theory- and replication-supported residual links, improving uncertainty quantification when full sampling covariance information becomes available, and extending the framework to joint genetic-clinical network models to evaluate whether residual hubs predict clinical trajectories or treatment outcomes beyond shared factor liability.

## Limitations

This was a secondary analysis based on published summary results. The joint sampling covariance across genetic correlations and SEM parameters was not available. The independence approximation could have underestimated uncertainty. The five factor plus p analysis used an operational baseline derived from available information. It may not reproduce the full specification of the original p factor model. Finally, heterogeneity in phenotype definitions, ascertainment, and comorbidity across GWAS cohorts, as well as the limited set of disorders available, may contribute to residual structure and limit generalizability.

## Conclusion

A five factor genetic structure captured broad shared liability but left structured residual overlap. Obsessive compulsive disorder and attention deficit hyperactivity disorder were consistent residual hubs across both within-domain and cross-domain links in the five-factor structure. These findings suggest that transdiagnostic genetic architectures are informative but incomplete. We encourage a reflective approach to psychiatric genetics in which large-scale data are used not only to confirm higher-order structure, but also to interrogate its limits and guide more explainable models of mental disorders.

## Data Availability

All data produced in the present study are available upon reasonable request to the authors.

